# Pandemic Lessons of Sustainability: Higher Covid-19 Mortality in Less Sustainable US States

**DOI:** 10.1101/2023.05.22.23290349

**Authors:** Lee Liu

## Abstract

This paper intends to contribute to the current debate over what lessons the United States should take away from the Covid-19 pandemic. It focuses on the role that sustainability played in shaping different pandemic outcomes among the 50 states. By the end of 2021, Mississippi reported the highest standardized death rate from Covid-19 in the country, more than five times higher than Vermont, which reported the lowest standardized death rate. If Mississippi had the same rate as Vermont, approximately 83% of the lives lost (7,958 individuals) could have been saved. If all 50 states had the same rate as Vermont, approximately 583,296 individuals (76% of the total deceased) would have survived. The inter-state difference in excess death rates was even larger. It was 18.19% in Arizona, 8.5 times as high as in Hawaii. Political ideology is currently a popular possible explanation for discrepancies among states in pandemic outcomes, given that Republican states tended to have higher death rates compared to Democratic ones. Additionally, partisan politics have been criticized for hindering the US pandemic response, especially in the early stages of the pandemic. However, the current debate lacks an attention to sustainability. This study demonstrates that indicators of sustainability may serve as more significant predictors of the death rates among the US states than political affiliation. Using the percentage of votes for Trump per state in 2020 as a proxy variable, this study found that the correlation between political affiliation and the death rates was significant only when it was the lone parameter. Its effects were overshadowed when vaccination rates and eco-friendliness were included in the equation. Above all, when the Sustainable Development Goal (SDG) index was added to the regression, it became the only significant predictor of the death rates. This suggests that it was not “red” or “blue,” but rather “green” that was the most important factor in determining Covid-19 mortality. Pandemic lessons are lessons of sustainability.

## Introduction

Numerous far-reaching publications have diligently investigated what the United States can learn from the Covid-19 pandemic. These include *Lessons from the Covid War* by a team of 34 experts (Covid Crisis Group, 2023), *World War C* (Gupta, 2021), and two Lancet papers (Sachs, et al., 2022; Bollyky, et al., 2023). Table 1 presents a list of selective lessons and factors affecting Covid-19 outcomes that have been described in these publications, including preparedness, institutions and leadership, partisan politics, public trust, inequalities, collaboration, public-private partnerships, scientific research, and communication. Notably, it has been reported that partisan politics hindered mitigation efforts. Republican states reported higher Covid-19 death rates as well as lower vaccination rates than Democratic states (Neelon et al., 2021; Krieger et al., 2022; Kaashoek et al., 2022; Wallace et al., 2022; Bolsena and Palmb, 2022). Such findings were also extensively reported in the national media (McPhillips, 2021; Mitropoulos, 2022; Denworth, 2022). The rapid development of vaccines has been lauded as evidence for the importance of scientific research and public-private partnership and collaboration. However, a sustainability perspective is lacking in the current debate. To contribute to this important undertaking, this paper examines why some US states were more impacted by the pandemic than others and what roles sustainability played in shaping outcomes of the pandemic.

**Table 1.** Selective Lessons/Factors in Covid Outcomes by Selective Publications

| Authors | Selective Lessons/Factors in Covid Outcomes |
| --- | --- |
| Covid Crisis Group, 2023 | Preparedness<br>Institutions and leadership<br>Inequalities<br>Public-private partnerships<br>Scientific research and collaboration<br>Communication |
| Bollyky, et al., 2023 | Social, racial, and economic inequities<br>Health disparities<br>Partisan politics<br>Public trust |
| Sachs, et al., 2022 | Investment in healthcare<br>Cooperation |
| WHO-SEARO, 2022 | Health emergency governance<br>Primary health care-oriented resilient health systems<br>Adaptable health-care system response with effective care pathways<br>Public-private partnerships<br>Risk communication and management of infodemics |
| Blau, et al., 2022 | Behavior change<br>Misinformation<br>Inequality |
| Gupta, 2021 | Early detection and response<br>Investing in healthcare infrastructure<br>Cooperation and coordination<br>Science and research<br>Public health education and communication<br>Preparedness and resilience |

Sustainability, or sustainable development, is a holistic approach to progress that aims to balance economic, social, and environmental considerations. To promote sustainable development and address key challenges, the UN Agenda 2030 was adopted by all UN member countries including the United States in 2015 (UN, 2015). The agenda consists of interconnected and interdependent Sustainable Development Goals (SDGs) to be achieved by the year 2030. The SDGs cover a wide range of issues that encompass all of the pandemic lessons either directly or indirectly. It has been reported that Asian countries with higher SDG indexes tended to have lower Covid-19 mortality rates (Zhou and Puthenkalam, 2022). Indeed, it is possible for sustainability to serve as an overarching theme in investigating key lessons from the pandemic. Current reports have focused on the pandemic’s negative impact on progress made towards achieving the SDGs by 2030. The pandemic has paused SDG implementation (UN 2020; UN/DESA 2020) and reversed progress made in poverty reduction, health and education, inequality, and basic services worldwide (UN, 2022; Lynch and Sachs, 2021). Furthermore, the pandemic necessarily diverted financial, human, and political resources away from sustainability efforts. However, it is important to note that the pandemic has also highlighted the importance of sustainability and resilience in the face of global crises. Pre-pandemic progress in achieving SDGs contributed to better mitigation and outcomes of the adverse impacts of the pandemic, while lack of progress towards achieving the SDGs can be seen as responsible for the severe impacts of the pandemic (UN/DESA 2020). Recovery from the pandemic presents an opportunity to rebuild a more sustainable and resilient world, if we can incorporate lessons learned from the crisis into future planning and development.

## Data and Methods

The 2020 and 2021 Covid-19 mortality data by state, adjusted for differences in age-distribution and population size, were obtained from the United States CDC (2023a). 2021 was the most recent yearly data available. The author derived death rates by the end of 2021 by using the CDC data on number of deaths and death rates in each year. Excess death rates can inform about the possible burden of deaths associated directly or indirectly with the pandemic (CDC, 2023b) and thus were used as an additional measurement of state Covid-19 mortality. The author derived the data from CDC’s weekly estimates of excess deaths associated with Covid-19 by the end of 2021 (CDC, 2023b). Post 2021 data were excluded because they were considered as incomplete. The CDC states that such data are incomplete as only 60% of the records have been submitted and “completeness varies by jurisdiction.” The formula used for calculation was: Excess death rate = (Observed deaths - Expected deaths) / Expected deaths. The CDC (2013b) has the following statement:

> Estimates of excess deaths can be calculated in a variety of ways, and will vary depending on the methodology and assumptions about how many deaths are expected to occur. Estimates of excess deaths presented in this webpage were calculated using Farrington surveillance algorithms (1). A range of values for the number of excess deaths was calculated as the difference between the observed count and one of two thresholds (either the average expected count or the upper bound of the 95% prediction interval), by week and jurisdiction.

State vaccination rates (%) for the completed primary series by the end of 2021 were accessed from the CDC (2023c). Voting data were from the 2020 National Popular Vote Tracker at the Cook Political Report with Amy Walter (Wasserman et al., 2023), and the percentage of votes for Trump in each state was used as a proxy of partisan politics. State eco-friendliness scores were from WalletHub. The scores (from 0 to 100) were based on rankings from 25 key metrics on environmental quality, eco-friendly behaviors, and climate change contributions (Kiernan, 2023). A score of 100 signifies the attainment of overall eco-friendliness. As such, eco-friendliness scores can be seen as a measure of a particular state’s progress towards sustainability. A more comprehensive measurement of sustainability, however, are the Sustainable Development Goal (SDG) index scores (from 0 to 100) published in the United States Sustainable Development Report 2021 by the UN Sustainable Development Solutions Network (SDSN) and SDSN USA (Lynch and Sachs, 2021). The SDG index is a tool used to measure and track progress towards the 17 UN SDGs with 169 targets and 232 unique indicators. A score of 100 indicates the achievement of all SDGs. The 2021 US Sustainable Development Report was based on data collected by the SDSN by the end of 2020. As such the scores reflected state achievement of SDGs before the pandemic.

Data analyses were conducted on the 50 US states, excluding Washington DC and other US territories, because the eco-friendliness and SDG index scores were only available at the state level. The SPSS software was used to derive descriptive statistics and correlation coefficients of all variables. Linear regression analyses were applied to investigate the relationship between the standardized Covid-19 death rates, excess death rates, and their possible explanatory variables.

## Results

Table 2 presents the descriptive statistics. The Covid-19 death rates ranged from 45.78 per 100,000 people in Vermont to 274.88 in Mississippi. This means that people living in Mississippi, the state with the highest Covid-19 death rate, were six times as likely to die of Covid as people in Vermont, the state with the lowest Covid-19 death rate. By the end of 2021, 9,548 Mississippians had died from Covid-19. If the state had had the same death rate as Vermont, 83% of them (7,958 Mississippians) would have lived. Meanwhile, 766,429 Americans across the 50 states died, with a national death rate of 191.56 per 100,000 population. If the death rate in all 50 states had been the same as that in Vermont, about 76% of the lives lost (583,296 people) by the end of 2021 would have been saved. The difference in excess death rates was even larger among the states. Arizona had the highest excess death rate at 18.19%, which was 8.5 times as high as the lowest rate at 2.13% in Hawaii. Total excess deaths in the United States exceeded 2.115 million people by the end of 2021, for which about 1.7 million would have lived if the country had had Hawaii’s rate. In the 2020 presidential election, the percentage of voters who voted for Trump ranged from 30.7% in Vermont to 69.9% in Wyoming. Difference in vaccination rates ranged from 49.1% in Alabama to 79.6% in Rhode Island. Vermont had the highest eco-friendliness scores (78.44%), three times higher than West Virginia (19.3%), which had the lowest scores. Vermont also had the highest reported 2021 SDG index (60%), which was twice the 2021 SDG index of Mississippi (30%).

**Table 2.** Descriptive statistics.

| Variable | Minimum | Maximum | Mean | Std. Deviation |
| --- | --- | --- | --- | --- |
| Standardized death rate<br>(per 100,000 population) | 45.78 | 274.88 | 182.39 | 51.82 |
| Excess death rate (%) | 2.13 | 18.19 | 10.85 | 3.8 |
| Votes for Trump (%) | 30.7 | 69.9 | 50.09 | 10.33 |
| Vaccination rate (%) | 49.1 | 79.6 | 62.14 | 8.53 |
| Eco-friendliness score | 19.30 | 78.44 | 57.03 | 12.97 |
| SDG index score | 30 | 60 | 45.50 | 8.14 |

N=50.

Pearson correlation results indicate that all variables were significantly correlated at the 99.5% level (Table 3). Higher Covid-19 death rates are associated with a higher percentage of votes for Trump, lower vaccination rates, lower eco-friendliness scores, and lower SDG index scores. Excess death rates were positively correlated to votes for Trump but negatively related to vaccination rates, eco-friendliness scores, and SDG index scores. Higher percentage of votes for Trump are associated with lower vaccination rates, eco-friendliness scores, and SDG index scores. Eco-friendliness was positively correlated with vaccination rate and SDG index.

**Table 3.** Pearson correlation (2-tailed) results.

|  | Standardized death rate | Excess death rate | Votes for Trump | Vaccination rate | Eco-friendliness |
| --- | --- | --- | --- | --- | --- |
| Excess death rate | 0.652** |  |  |  |  |
| Votes for Trump | 0.553** | 0.390* |  |  |  |
| Vaccination | -0.665** | -0.577** | -0.869** |  |  |
| Eco-friendliness | -0.616** | -0.603** | -0.801** | 0.756** |  |
| SDG index | -0.756** | -0.664** | -0.755** | 0.807** | 0.883** |

\* indicates significance at 99.5%.
\*\* indicates significance at 99.9%.

Table 4 displays the progression of regression models used for state Covid-19 death rates. The explanatory variables included votes for Trump, vaccination rate, eco-friendliness, and SDG index. Regression A used votes for Trump (%) as the single variable, which proved statistically significant and could explain about 30% of the variations in death rates among states. Vaccination rate (%) replaced votes for Trump as the primary explanatory factor when it was added to the equation (B), resulting in a higher explanatory value. Both vaccination rate and eco-friendliness score, but not votes for Trump, had statistically significant effects on the death rates when all three variables were included in Regression C. SDG index was the only statistically significant explanatory factor when it entered the equation (D). The effects of vaccination and eco-friendliness were no longer significant. Votes for Trump had no statistically significant effect on the death rate when it was used with any of the other three variables. On the other hand, SDG index was consistently the only explanatory factor with a significance of 99.9% (D–G). It was able to account for over 55% of the changes in the death rates in all regression models (D–G) (Figure 1).

**Table 4.** Regression results for US state standardized Covid-19 death rates.

|  | A | B | C | D | E | F | G |
| --- | --- | --- | --- | --- | --- | --- | --- |
| Constant | 45.378<br>(30.833) | 490.333**<br>(138.933) | 595.733***<br>(143.272) | 525.402***<br>(126.046) | 389.687***<br>(78.228) | 565.089***<br>(116.962) | 420.394***<br>(73.811) |
| Votes for Trump | 2.775***<br>(0.603) | -0.495<br>(1.102) | -1.593<br>(1.185) | -0.805<br>(1.088) | 0.202<br>(0.968) | -1.229<br>(0.987) | -0.203<br>(0.729) |
| Vaccination |  | -4.556***<br>(1.335) | -4**<br>(1.317) | -1.82<br>(1.33) |  | -2.055<br>(1.301) |  |
| Eco-friendliness |  |  | -1.49*<br>(0.716) | 0.783<br>(0.911) | 1.038<br>(0.9) |  |  |
| SDG index |  |  |  | -5.148***<br>(1.466) | -6.079***<br>(1.309) | -4.252***<br>(1.029) | -5.007***<br>(0.925) |
| R<sup>2</sup> | 0.306 | 0.444 | 0.492 | 0.601 | 0.585 | 0.595 | 0.573 |
| Adjusted R<sup>2</sup> | 0.292 | 0.42 | 0.459 | 0.566 | 0.558 | 0.568 | 0.554 |
| F | 21.175 | 18.762 | 14.839 | 16.955 | 21.58 | 22.487 | 31.485 |
| p | <0.001 | <0.001 | <0.001 | <0.001 | <0.001 | <0.001 | <0.001 |
| N | 50 |  |  |  |  |  |  |

Standard errors are reported in parentheses.
\* indicates significance at 95%.
\*\* indicates significance at 99%.
\*\*\* indicates significance at 99.9%.

**Figure 1.**
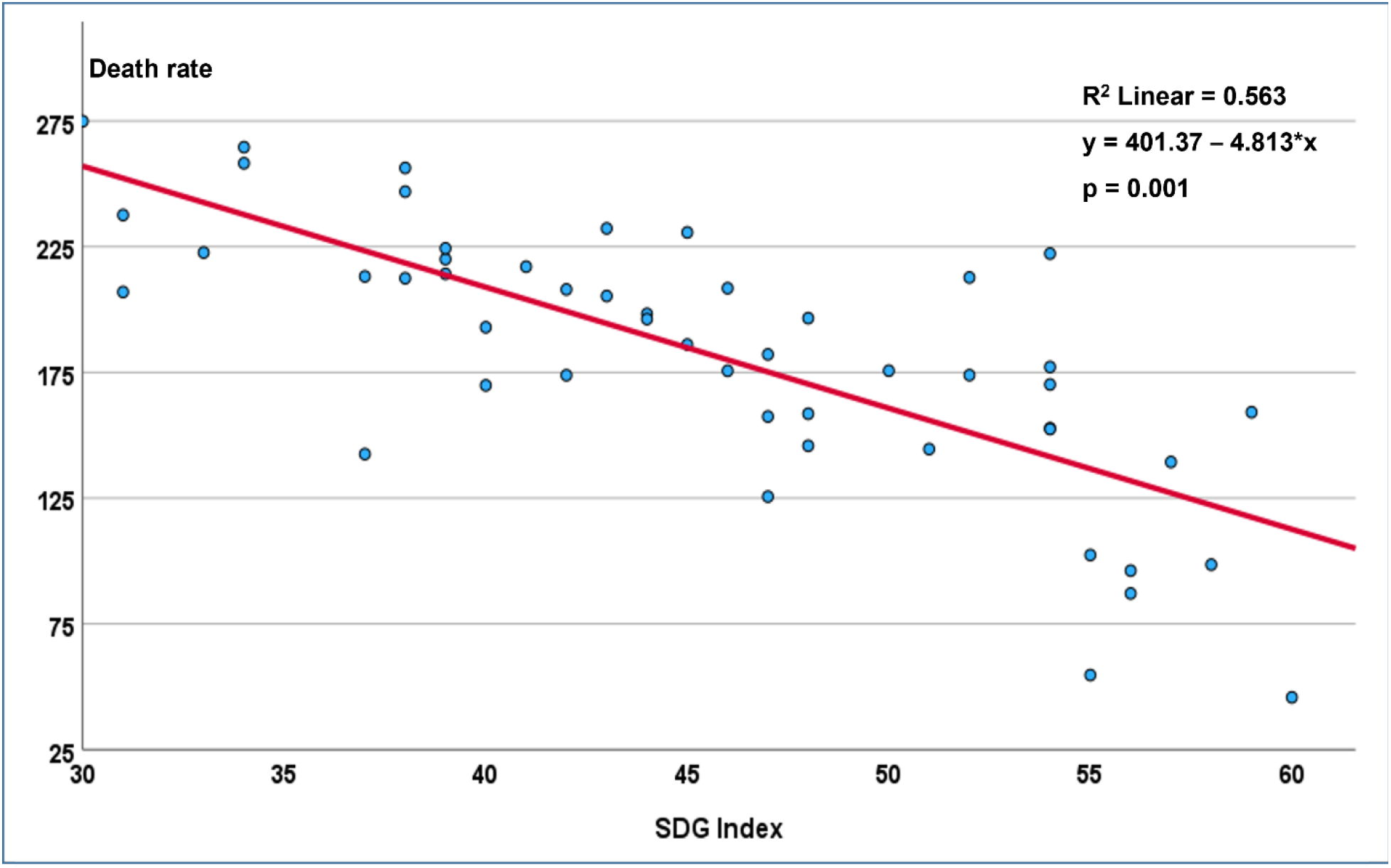
Relationship between standardized Covid-19 death rate per 100,000 and Sustainable Development Goal (SDG) index score among the 50 US states.

The progression of regression for state excess death rates is shown in Table 5. Regression A used votes for Trump as the single variable, which was statistically significant. Vaccination rate became the primary explanatory factor in Regression B. The effect of eco-friendliness score was statistically significant in Regression C. SDG index was the only statistically significant explanatory factor in Regressions D–G. SDG index was consistently the only explanatory factor with a significance of 95% (D–G). It was able to explain 44% of the changes in the excess death rates (Figure 2).

**Table 5.** Regression results for US state Covid-19 associated excess death rates.

|  | A | B | C | D | E | F | G |
| --- | --- | --- | --- | --- | --- | --- | --- |
| Constant | 3.652<br>(2.502) | 46.142***<br>(10.35) | 25.201***<br>(3.248) | 36.286***<br>(6.406) | 24.801***<br>(2.401) | 33.801***<br>(6.042) | 26.219***<br>(3.044) |
| Votes for Trump | 1.44**<br>(0.049) | -0.167<br>(0.085) |  | -0.217<br>(0.066) |  | -0.094<br>(0.06) |  |
| Vaccination |  | -0.433***<br>(0.103) | -1.26<br>(0.77) |  |  |  | -0.053<br>(0.082) |
| Eco-friendliness |  |  | -0.114*<br>(0.051) | -0.084<br>(0.074) | -0.022<br>(0.068) |  |  |
| SDG index |  |  |  | -0.084**<br>(0.074) | -0.279*<br>(0.109) | -0.4***<br>(0.076) | -0.265***<br>(0.086) |
| R<sup>2</sup> | 0.152 | 0.383 | 0.398 | 0.483 | 0.441 | 0.469 | 0.445 |
| Adjusted R<sup>2</sup> | 0.135 | 0.357 | 0.372 | 0.449 | 0.418 | 0.466 | 0.422 |
| F | 8.627 | 14.617 | 15.517 | 14.333 | 18.577 | 20.72 | 18.861 |
| p | <0.005 | <0.001 | <0.001 | <0.001 | <0.001 | <0.001 | <0.001 |
| N | 50 |  |  |  |  |  |  |

Standard errors are reported in parentheses.
\* indicates significance at 95%.
\*\* indicates significance at 99.5%.
\*\*\* indicates significance at 99.9%.

**Figure 2.**
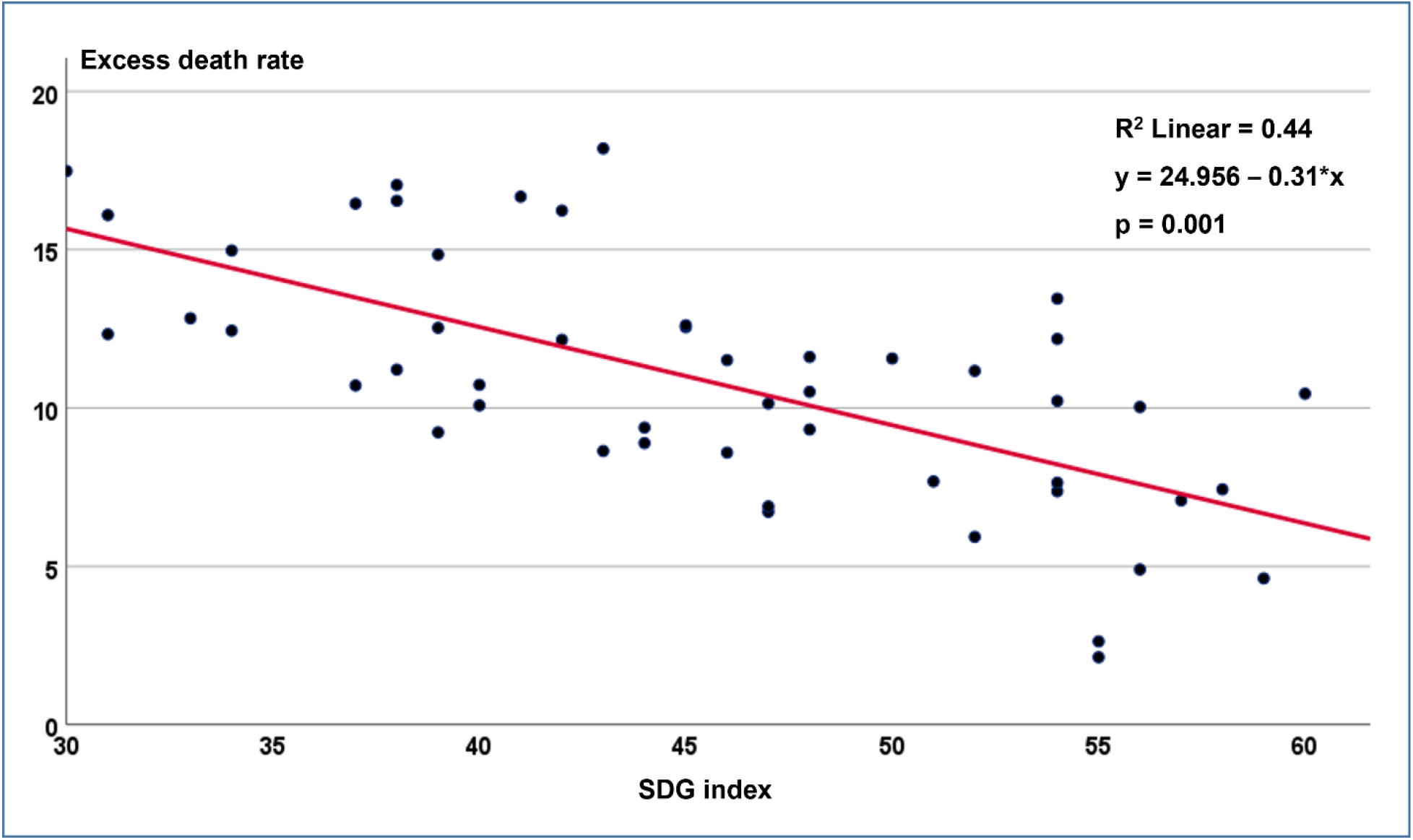
Relationship between Covid-19 associated excess death rate (%) and Sustainable Development Goal (SDG) index score among the 50 US states.

As expected, state vaccination rates were negatively affected by votes for Trump, which appears to be a dominant factor (Table 6). The positive effect of SDG index was also significant but seemingly less influential than votes for Trump. The model was able to explain over 80% of variations in vaccination rates among the states. The effects of eco-friendliness were insignificant.

**Table 6.** Regression results for US state vaccination rates (Completed primary series by the end of 2021).

|  |  |
| --- | --- |
| Constant | 83.071***<br>(10.536) |
| Votes for Trump | −0.549***<br>(0.088) |
| Eco-friendliness | −0.117<br>(0.99) |
| Death rates | −0.22<br>(0.016) |
| SDG index | 0.379**<br>(0.172) |
| R<sup>2</sup> | 0.823 |
| Adjusted R<sup>2</sup> | 0.808 |
| F | 52.453 |
| p | <0.001 |
| N | 50 |

Standard errors are reported in parentheses.
\*\* indicates significance at 95%.
\*\*\* indicates significance at 99.9%.

## Discussion

Partisan politics was regarded as one of the key factors weakening the United States’ ability to fight the pandemic (Table 1). On the other hand, the United States has been praised for its scientific research and public-private partnership (Covid Crisis Group, 2023), which led to the rapid development of vaccines (Operation Warp Speed). The findings here indicate that there are additional lessons to be learned. Votes for Trump, as a proxy factor for partisan politics, affected standardized and excess death rates, but became insignificant when other factors were included in the equation. Instead, vaccination and eco-friendliness became the primary explanatory factors. Their effects became insignificant when SDG index was added to the equation. Such changes in effectiveness may be because of confounding. Confounding occurs when the relationship between the dependent variable and one independent variable becomes distorted or obscured by the presence of another independent variable. It should be noted that eco-friendliness scores are based on environmental quality, environment-friendly behaviors, and climate-change contribution, which belong to the environmental component of sustainability. They affect health outcomes, including Covid-19 death rates. However, it is understandable that their effects may be overshadowed by the effect of SDG index scores.

In terms of standardized death rates, eco-friendliness was more important than partisan politics and nearly as important as vaccination in terms of the explanatory power. It was also more important than vaccination in explaining variation in state excess death rates. Most notably, the findings of this report suggest that sustainability, measured as pre-pandemic SDG index scores, might have a powerful effect on both standardized and excess death rates.

Vaccination has been regarded as the most effective preventative measure against infectious diseases (Andre et al., 2008) and vaccination was a key player in the United States’ fight against the pandemic (Covid Crisis Group, 2023). The findings suggest that partisan politics severely limited vaccination rates in certain states, however, while indicators of sustainability had a positive effect on vaccination.

In fact, the lessons described in the publications (Table 1) are ultimately lessons of sustainability as explained below. The following discussion also helps to understand why sustainability was the key factor in the Covid war.

### Preparedness

The pandemic revealed serious deficiencies in the US health care systems (Covid Crisis Group, 2023; Gupta, 2021; Blumenthal et al., 2020; Dorsett, 2020). The US lacks capacity in primary care: Covid-19 mitigation was hindered by the lack of healthcare insurance coverages, the financial loss to healthcare facilities and professionals, racial and ethnic disparities in healthcare coverages, and lack of public healthcare capacity (Blumenthal et al., 2020). Care was delayed among the earliest infections of Covid-19, further accelerating outbreaks due to the unpreparedness of the healthcare system, including high costs of healthcare in general (Hook and Kuchler, 2020) and lack of access to healthcare providers, as many Americans are uninsured or underinsured (Blumenthal and Seervai, 2020). Had the United States achieved implementation of SDG 3, which requires “strengthening healthcare system capacity for early warning, risk reduction and management of health risks” to “ensure healthy lives and promote well-being for all” (UN, 2015), the country would have been better prepared to deal with the Covid-19 crisis. SDG 3 also demands quality healthcare and “universal health coverage (UN, 2015),” which would have curbed outbreaks among those who were delayed care because of financial concerns.

### Institutions, Leadership, Partisan Politics, and Public Trust

The US fought the Covid War with a 19^th^ century institutional system (Covid Crisis Group, 2023). Partisan politics reduced public trust in government (Covid Crisis Group, 2023; Bollyky, et al., 2023). This may have contributed to a slower recognition and response to the pandemic, which worsened outbreaks in the US (Covid Crisis Group, 2023; Bollyky, et al., 2023; Gupta, 2021; Schneider 2020; Diamond and Wheaton 2020). Global and national leadership could have effectively prevented the outbreaks from becoming a pandemic (Covid Crisis Group, 2023; Bollyky, et al., 2023; Blau, et al., 2022; WHO-SEARO, 2022). Unfortunately, such leadership was not present, along with a lack of international and national cooperation and coordination. For instance, if the US federal government had coordinated a national exchange of ventilators among its states, it would have been able to reduce American deaths by the thousands (Adelman, 2020). These challenges would have been better met if SDG 16, which calls for “effective, accountable and transparent institutions” had been achieved (UN, 2015).

### Inequalities

The pandemic disproportionately affected vulnerable communities in the United States (Covid Crisis Group, 2023; Bollyky, et al., 2023; Blau, et al., 2022; Lynch and Sachs, 2021; Maroko et al., 2020; Karaye and Horney, 2020). Such hotspots could have been minimized by achieving SDG 3 for health equity, SDG 10 for reducing income inequalities, SDG 1 on poverty elimination, and SDG 11 for developing inclusive cities (UN, 2015).

### Collaboration, Public-Private Partnerships

Many successful cases, such as vaccine development, and many failures were discussed in the literature (Covid Crisis Group, 2023; Sachs, et al., 2022; WHO-SEARO, 2022; Gupta, 2021). The SDGs emphasize the importance of collaboration among governments, NGOs, civil society, and the private sector (UN, 2015). Cooperation involves sharing knowledge, resources, and best practices to tackle common challenges, such as poverty, inequality, and climate change. The SDGs recognize the need for inclusive partnerships that involve diverse actors. Collaboration between governments, businesses, NGOs, academia, and communities enhances innovation, capacity-building, and the mobilization of resources. Cooperation, coordination, and public-private partnership are crucial aspects of achieving the SDGs (UN, 2015).

### Science and Research

It is widely agreed that scientific research was key to fighting the Covid-19 pandemic (Covid Crisis Group, 2023; WHO-SEARO, 2022; Gupta, 2021). The SDGs encompass a wide range of challenges that societies must apply scientific research and innovations to address (UN, 2015). For example, SDG 3, good health and well-being, depends on scientific research to advance medical knowledge, develop new treatments and vaccines, improve healthcare systems, and contribute to disease prevention and health promotion.

### Communication

Communication is crucial in fighting disinformation (Covid Crisis Group, 2023; Bollyky, et al., 2023; Blau, et al., 2022; WHO-SEARO, 2022). Governments and social media have played an increasing role in spreading fake news about Covid-19 and vaccines, causing confusion and disinformation, and making it difficult to protect the people. Xenophobia and discrimination both worsened Covid-19 outbreaks and were exacerbated by the pandemic in turn (Human Rights Watch, 2020; Gover et al., 2020). For example, anti-Asian sentiment and crimes increased in the United States following the first cases of Covid-19 in the country (Gover et al., 2020; Gao and Liu, 2021). Politicians and media outlets with extreme anti-diversity and xenophobic views played an important role in fanning the anti-Asian flames during the pandemic (Li and Nicholson, 2021). The achievement of at least two different SDGs would have directly contributed to the mitigation of these problems, such as SDG 11 for inclusive societies and SDG 16 for strong institutions (UN, 2015). For instance, pre-pandemic pro-diversity attitudes in urban Germany were shown to have prevented prejudice and discrimination during the pandemic (Drouhot, 2021). More effective communication of sustainability values would have made a difference in pandemic outcomes.

## Limitations and Further Research

The paper has several limitations. First, the data used in the statistical analyses have limitations. For the SDG index scores, some indicators had to measured using data reported a few years earlier as more recent data were unavailable; some were not measured at all because of lack of data or funding (Lynch and Sachs, 2021). Second, excess death rates were estimates only and subject to differences due to differences in methods of estimation, as stated in the Data and Methods section. Third, there are possibly other significant predictors of Covid-19 death rates for which data were unavailable and thus excluded from the analyses. Last, a strong correlation between two variables does not necessarily imply a cause-and-effect relationship. While a strong correlation may suggest a possible causal connection, there could be other factors or explanations that contribute to the observed relationship. These limitations mean that caution is required when interpreting the findings and it is important to conduct further research and consider other evidence to establish a causal relationship.

## Conclusions

Partisan politics has been publicized as a main factor contributing to differences in Covid-19 death rates throughout the United States. Additionally, science and technology in tandem with public-private partnership made the expedited development of vaccines possible. Vaccines are the most effective method for disease control if the population is willing to use them, and this study suggests that percentage of votes for Trump was one of the main variables in predicting vaccination rates. However, in addition to vaccination, the adoption of sustainable practices and the achievement of the SDGs will allow societies to be more prepared and resilient to crises like the Covid-19 pandemic in the long run. It is not Republicans or Democrats, but sustainability that is the key predictor in explaining the variations in Covid-19 death rates among the 50 US states. Vermont has the highest SDG index score and the lowest Covid-19 death rate among US states, despite having a Republican governor. Ultimately, lessons from the pandemic are lessons of sustainability. To become better prepared for the next pandemic or climate emergency, it is crucial for societies to adopt sustainable development principles and achieve sustainability overall.

## Data Availability

The research was based on publicly available data, as stated in the Data and Methods section.

https://www.cdc.gov/nchs/pressroom/sosmap/covid19_mortality_final/COVID19.htm

https://covid.cdc.gov/covid-data-tracker

https://us-states.sdgindex.org/downloads

## Acknowledgements

The author thanks Tiffany Liu for editing the paper

## Data Availability Statement

The research was based on publicly available data, as stated in the Data and Methods section.

## References

Adelman D. 2020. Thousands of lives could be saved in the US during the Covid-19 pandemic if states exchanged ventilators. Health Affairs, 39, 1247–125 DOI:10.1377/hlthaff.2020.00505

Andre FE, Booy R, Bock HL, et al. 2008. Vaccination greatly reduces disease, disability, death and inequity worldwide. Bull World Health Organ. 86: 140–146. doi: 10.2471/BLT.07.040089

Blau W, Bond H, Crete E, et al. 2022. Lessons from COVID-19 for Climate Change. UN SDSN White Paper. https://resources.unsdsn.org/science-for-a-sustainable-future.

Blumenthal D, Seervai S. 2020. Coronavirus Is Exposing Deficiencies in U.S. Health Care. Harvard Business Review. March 10, 2020. https://hbr.org/2020/03/coronavirus-is-exposing-deficiencies-in-u-s-health-care

Blumenthal, D, Fowler, EJ, Melinda Abrams, and Collins, SR. 2020. Covid-19 — Implications for the Health Care System. N Engl J Med 2020; 383:1483–1488. DOI: 10.1056/NEJMsb2021088

Bollyky TJ, Castro E, Aravkin AY, Bhangdia K, Dalos J, Hulland EN, et al. 2023. Assessing COVID-19 pandemic policies and behaviours and their economic and educational trade-offs across US states from Jan 1, 2020, to July 31, 2022: an observational analysis. Lancet 401(10385):1341–60.

Bolsena T and Palmb R. 2022. Politicization and COVID-19 vaccine resistance in the U.S. Prog Mol Biol Transl Sci. 2022; 188: 81–100. doi: 10.1016/bs.pmbts.2021.10.002

CDC (Centers for Disease Control and Prevention). 2023a. COVID-19 Mortality by State (Last reviewed: February 15, 2023). Accessed May 12, 2023, athttps://www.cdc.gov/nchs/pressroom/sosmap/covid19_mortality_final/COVID19.htm.

CDC. 2023b. National and State Estimates of Excess Deaths, Excess Deaths Associated with COVID-19. https://www.cdc.gov/nchs/nvss/vsrr/covid19/excess_deaths.htm

CDC. 2023c. COVID-19 Vaccinations in the United States. COVID Data Tracker. Atlanta, GA: US Department of Health and Human Services, CDC; Accessed on May 19, 2023, at https://covid.cdc.gov/covid-data-tracker

Covid Crisis Group. 2023. Lessons from the Covid War: An Investigative Report. Public Affairs.

Denworth L. 2022. People in Republican Counties Have Higher Death Rates Than Those in Democratic Counties. Scientific America, July 18, https://www.scientificamerican.com/article/people-in-republican-counties-have-higher-death-rates-than-those-in-democratic-counties/

Diamond D and Wheaton S. 2020. ‘The U.S. has hamstrung itself’: How America became the new Italy on coronavirus. Politico, 06/22/2020. https://www.politico.com/news/2020/06/22/united-states-italy-traded-places-coronavirus-333122

Dorsett M. 2020. Point of no return: COVID-19 and the U.S. healthcare system: An emergency physician’s perspective. Science Advances, 6 (26), eabc5354. DOI: 10.1126/sciadv.abc5354

Drouhot LG, Petermann S, Schönwälder K & Vertovec S. 2021. Has the Covid-19 pandemic undermined public support for a diverse society? Evidence from a natural experiment in Germany. Ethnic and Racial Studies, 44: 877–892, DOI: 10.1080/01419870.2020.1832698

Gao Q and Liu XF. 2021. Stand against anti-Asian racial discrimination during COVID-19: A call for action. International Social Work, 64: 261–264. DOI: 10.1177/0020872820970610

Gover AR, Harper SB, Langton L. 2020. Anti-Asian Hate Crime During the COVID-19 Pandemic: Exploring the Reproduction of Inequality. American Journal of Criminal Justice, 45: 647–667. DOI: 10.1007/s12103-020-09545-1

Gupta S. 2021. World War C: Lessons from the COVID-19 Pandemic and How to Prepare for the Next One. Simon & Schuster.

Hook L. Kuchler H. 2020. How coronavirus broke America’s healthcare system. FT Magazine, April 29, https://www.ft.com/content/3bbb4f7c-890e-11ea-a01c-a28a3e3fbd33

Human Rights Watch. 2020. Covid-19 Fueling Anti-Asian Racism and Xenophobia Worldwide. May 12, https://www.hrw.org/news/2020/05/12/covid-19-fueling-anti-asian-racism-and-xenophobia-worldwide

Kaashoek J, Testa C, Chen JT, Stolerman LM, Krieger N, Hanage WP, et al. (2022) The evolving roles of US political partisanship and social vulnerability in the COVID-19 pandemic from February 2020–February 2021. PLOS Glob Public Health 2(12): e0000557. https://doi.org/10.1371/journal.pgph.0000557

Karaye IM, Horney JA. 2020. Impact of Social Vulnerability on COVID-19 in the U.S.: An Analysis of Spatially Varying Relationships. American Journal of Preventive Medicine, 59, 317–325, September 01, 2020. DOI:https://doi.org/10.1016/j.amepre.2020.06.006

Kiernan JS. 2023. 2023’s Greenest States. April 2, 2023. WalletHub.https://wallethub.com/edu/greenest-states/11987

Krieger N, Testa C, Chen JT, Hanage WP, McGregor AJ. 2022. Relationship of political ideology of US federal and state elected officials and key COVID pandemic outcomes following vaccine rollout to adults: April 2021–March 2022. The Lancet Regional Health, 16. DOI: https://doi.org/10.1016/j.lana.2022.100384

Li Y. and Nicholson H.L. Jr. 2021. When “model minorities” become “yellow peril”—Othering and the racialization of Asian Americans in the COVID-19 pandemic. Sociology Compass, 15: e12849

Lynch A, Sachs J. 2021. The United States Sustainable Development Report 2021. NewYork: SDSN. Accessed May 12, 2023, at https://us-states.sdgindex.org/downloads

Maroko, AR, Nash, D, Pavilonis, BT. 2020. COVID-19 and Inequity: a Comparative Spatial Analysis of New York City and Chicago Hot Spots. J Urban Health. 2020 Aug; 97: 461–470. DOI: 10.1007/s11524-020-00468-0

McPhillips D. 2021. Covid case and death rates were higher in GOP-led states in second half of 2020, study finds. CNN. March 10. https://www.cnn.com/2021/03/10/politics/covid-cases-deaths-red-blue-states-late-2020/index.html

Mitropoulos A. 2022. For red and blue America, a glaring divide in COVID-19 death rates persists 2 years later. ABC News. March 28, https://abcnews.go.com/Health/red-blue-america-glaring-divide-covid-19-death/story?id=83649085.

Neelon B, Mutiso F, Mueller NT, Pearce JL, Benjamin-Neelon SE. 2021. Associations Between Governor Political Affiliation and COVID-19 Cases, Deaths, and Testing in the U.S. American Journal of Preventive Medicine, 61:115–119. doi:10.1016/j.amepre.2021.01.034

Sachs JD, Karim SSA, Aknin L, Allen J, Brosbøl K, Colombo F, et al. 2022. The Lancet Commission on lessons for the future from the COVID-19 pandemic. Lancet, 400:1224–80.

Schneider EC. 2020. Failing the test — the tragic data gap undermining the U.S. pandemic response. N Engl J Med. 383: 299–302.

Spotswood, E.N., Benjamin, M., Stoneburner, L. et al. 2021. Nature inequity and higher COVID-19 case rates in less-green neighbourhoods in the United States. Nat Sustain, 4: 1092–1098. https://doi.org/10.1038/s41893-021-00781-9

UN. 2022. The Sustainable Development Goals Report 2022.https://www.un.org/sustainabledevelopment/progress-report/

UN. 2015. Transforming our world: the 2030 Agenda for Sustainable Development. UnitedNations, https://sustainabledevelopment.un.org/post2015/transformingourworld

UN/DESA. 2020. UN/DESA Policy Brief #78: Achieving the SDGs through the COVID-19response and recovery. United Nations. 11 June 2020https://www.un.org/development/desa/dpad/publication/un-desa-policy-brief-78-achieving-the-sdgs-through-the-covid-19-response-and-recovery/

UNDP (United Nations Development Programme). 2020. UN sets out COVID-19 social and economic recovery plan. UN, April 27.https://www.undp.org/content/undp/en/home/news-centre/news/2020/UN_sets_out_COVID_social_and_economic_recovery_plan.html

Wallace J, Goldsmith-Pinkham P, Schwartz JL. 2022. Excess Death Rates for Republicans and Democrats During the COVID-19 Pandemic. NBER (National Bureau of Economic Research) Working Paper No. 30512 September 2022.S

Wasserman D, Andrews S, Saenger L, Cohen L, Flinn A, and Tatarsky G. 2023. 2020National Popular Vote Tracker. The Cook Political Report with Amy Walter. Accessed May 12, 2023, at https://www.cookpolitical.com/2020-national-popular-vote-tracker

WHO-SEARO (World Health Organization Regional Office for South-EastAsia). 2022. Lessons learned from COVID-19 pandemic: virtual regional consultation with informal expert group. World HealthOrganization. https://apps.who.int/iris/handle/10665/363472.

Zhou L, Puthenkalam JJ. 2022. Correlation of the sustainable development goals index score and COVID-19 death rate: a comparison among 40 Asian countries, International Journal of Sustainable Development & World Ecology, 29:8, 840–849, DOI: 10.1080/13504509.2022.2107107

